# Evaluating Data-Driven Forecasting Methods for Predicting SARS-CoV2 Cases: Evidence From 173 Countries

**DOI:** 10.1101/2020.08.03.20167189

**Authors:** Ghufran Ahmad, Furqan Ahmed, Suhail Rizwan, Javed Muhammad, Hira Fatima, Aamer Ikram, Hajo Zeeb

**Affiliations:** National University of Sciences and Technology (NUST), Islamabad, Pakistan; Leibniz Institute for Prevention Research and Epidemiology, Bremen, Germany; University of Swabi, Pakistan; University of Adelaide, Australia; National Institute of Health (NIH), Pakistan; Health Sciences Bremen, University of Bremen, Bremen, Germany

## Abstract

The WHO announced the epidemic of SARS-CoV2 as a public health emergency of international concern on 30th January 2020. To date, it has spread to more than 200 countries, and has been declared as a global pandemic. For appropriate preparedness, containment, and mitigation response, the stakeholders and policymakers require prior guidance on the propagation of SARS-CoV2. This study aims to provide such guidance by forecasting the cumulative COVID-19 cases up to 4 weeks ahead for 173 countries, using four data-driven methodologies; autoregressive integrated moving average (*ARIMA*), exponential smoothing model (*ETS*), random walk forecasts (*RWF*) with and without drift. We also evaluate the accuracy of these forecasts using the Mean Absolute Percentage Error (MAPE). The results show that the *ARIMA* and *ETS* methods outperform the other two forecasting methods. Additionally, using these forecasts, we generated heat maps to provide a pictorial representation of the countries at risk of having an increase in cases in the coming 4 weeks for June. Due to limited data availability during the ongoing pandemic, less data-hungry forecasting models like *ARIMA* and *ETS* can help in anticipating the future burden of SARS-CoV2 on healthcare systems.

## Introduction

Severe Acute Respiratory Syndrome Coronavirus-2 (SARS-CoV2) is a zoonotic virus belonging to the betacoronavirus group of the coronaviridae family which also includes SARS-CoV and MERS. These viruses are known to cause severe acute respiratory diseases in humans.^1^ The first confirmed case of SARS-CoV2 emerged in the Wuhan city of the Hubei province, China in December 2019. The WHO announced the epidemic of SARS-CoV2 as a public health emergency of international concern on 30th January 2020 due to the high human to human transmission rate and absence of any treatment or vaccine.^2^ To date, it has spread to more than 200 countries and has been declared as a global pandemic.^3^ SARS-CoV2 transmits through respiratory droplets and has a binding capacity, through spike proteins, to angiotensin-converting enzyme 2 (ACE2) receptors in the human respiratory system.^3^ Clinical symptoms of SARS-CoV2 include cough, fever, shortness of breath, and – in severe cases – pneumonia and multiple organ failure.^3^ SARS-CoV2 has an incubation period of 1-14 days and a substantial proportion of the infected persons appear to be asymptomatic. Moreover, these individuals are highly infectious before the onset of symptoms, which makes it a challenge to diagnose, contain and control transmissions.^1,4^ Initially, the mean basic reproduction number (*R*_0_) of SARS-CoV2 ranged from 1.4 to 6.49, while some studies highlighted that the *R*_0_ stabilized around 2-3 leading to an exponential increase in the number of cases.^3,5^

Globally, many public and private enterprises are exploring treatment options and are in the process of vaccine development. However, vaccines have to go through a robust and usually time-consuming process of clinical trials owing to the paradigms of human safety, health, and bioethics.^6^ Some experts have shown concern that a vaccine for SARS-CoV2 may never even be developed and, even if developed, its production could take 1-2 years.^7^ Mutations of the virus are also a major concern in this regard since the virus has shown 14 mutations from December 2019 to March 23, 2020.^8^ Globally, countries have implemented various interventions in an attempt to limit transmissions and curtail the number of deaths caused by COVID-19. These interventions include social distancing, the closing of public places, academic institutes and schools, travel restrictions, quarantine for the infected, and – in some cases – curfew or lockdown.^9^ However, issues in the health security infrastructure, disease surveillance, health systems, and limited availability of health professionals, makes it a challenge for containing and mitigating infectious outbreaks.^9^

Considering these concerns, for appropriate preparedness, containment, and mitigation response, the stakeholders and policymakers require prior guidance on the propagation of SARS-CoV2. This study aims to provide such guidance by forecasting the cumulative COVID-19 cases up to 4 weeks ahead, and ascertain their accuracy using the Mean Absolute Percentage Error.

## Data Sources

The daily level data for the cumulative COVID-19 cases, for 173 countries, and at the aggregated level for the entire world, was acquired from “Our World in Data” – a combined effort of the researchers at the University of Oxford and the Global Change Data Lab – which relies on the European Centre for Disease Prevention and Control (ECDC) for data collection.^10,11^ Our sample period starts from the day of the first reported case for each country till 1^st^ June 2020. The statistical analysis of this was performed using R 3.6.3.

Our variable of interest, for the forecasting analysis, is the cumulative COVID-19 cases at the daily level. The descriptive statistics – number of observations, standard deviation, minimum, and maximum – are presented in Table 1. For brevity, Table 1 only provides the descriptive statistics for the 29 countries with the highest cumulative COVID-19 cases as of 1^st^ June 2020 and at the aggregated level for the entire world. The descriptive statistics for the 173 countries are provided in Table 1 of the Online Appendix.

**Table 1:**
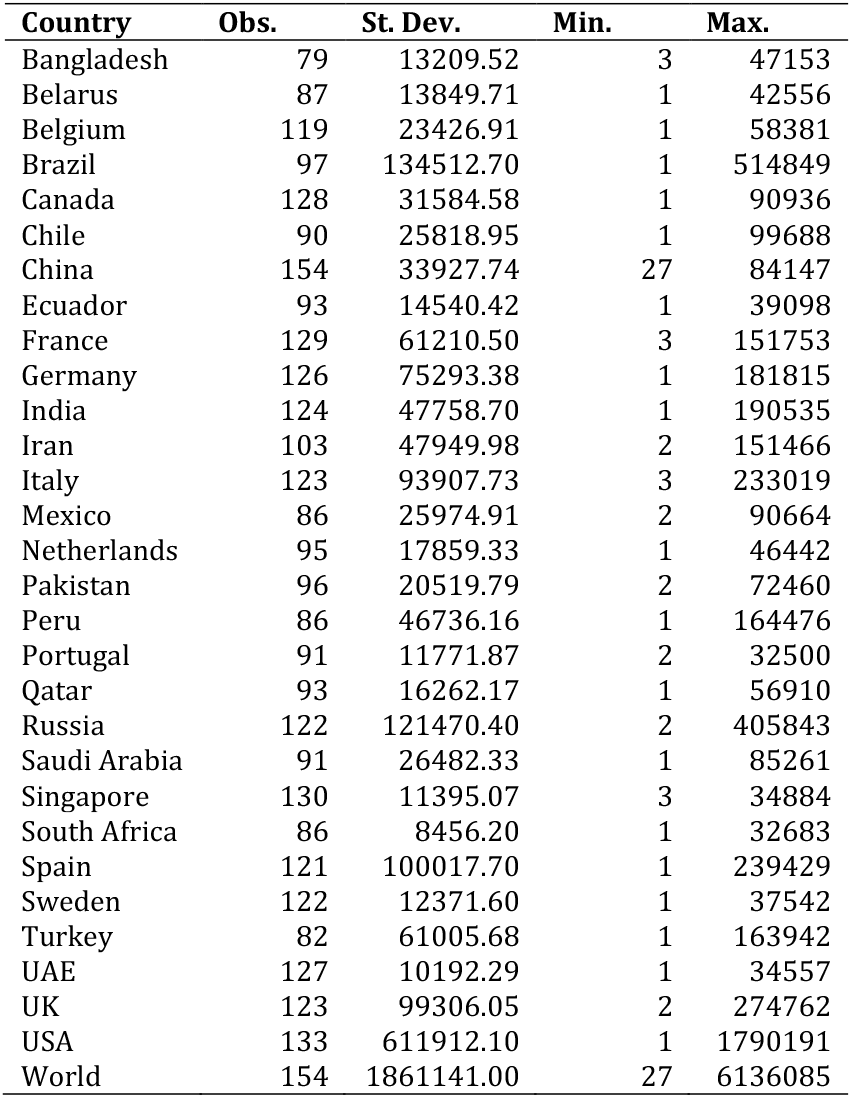
Descriptive statistics of the 29 countries with the highest cumulative COVID-19 cases and at the aggregated-level of the entire world.

| Country | Obs. | St. Dev. | Min. | Max. |
| --- | --- | --- | --- | --- |
| Bangladesh | 79 | 13209.52 | 3 | 47153 |
| Belarus | 87 | 13849.71 | 1 | 42556 |
| Belgium | 119 | 23426.91 | 1 | 58381 |
| Brazil | 97 | 134512.70 | 1 | 514849 |
| Canada | 128 | 31584.58 | 1 | 90936 |
| Chile | 90 | 25818.95 | 1 | 99688 |
| China | 154 | 33927.74 | 27 | 84147 |
| Ecuador | 93 | 14540.42 | 1 | 39098 |
| France | 129 | 61210.50 | 3 | 151753 |
| Germany | 126 | 75293.38 | 1 | 181815 |
| India | 124 | 47758.70 | 1 | 190535 |
| Iran | 103 | 47949.98 | 2 | 151466 |
| Italy | 123 | 93907.73 | 3 | 233019 |
| Mexico | 86 | 25974.91 | 2 | 90664 |
| Netherlands | 95 | 17859.33 | 1 | 46442 |
| Pakistan | 96 | 20519.79 | 2 | 72460 |
| Peru | 86 | 46736.16 | 1 | 164476 |
| Portugal | 91 | 11771.87 | 2 | 32500 |
| Qatar | 93 | 16262.17 | 1 | 56910 |
| Russia | 122 | 121470.40 | 2 | 405843 |
| Saudi Arabia | 91 | 26482.33 | 1 | 85261 |
| Singapore | 130 | 11395.07 | 3 | 34884 |
| South Africa | 86 | 8456.20 | 1 | 32683 |
| Spain | 121 | 100017.70 | 1 | 239429 |
| Sweden | 122 | 12371.60 | 1 | 37542 |
| Turkey | 82 | 61005.68 | 1 | 163942 |
| UAE | 127 | 10192.29 | 1 | 34557 |
| UK | 123 | 99306.05 | 2 | 274762 |
| USA | 133 | 611912.10 | 1 | 1790191 |
| World | 154 | 1861141.00 | 27 | 6136085 |

## Ethics

No ethics approval was required for the study as secondary data analysis was performed on publicly available COVID-19 dataset.

## Forecasting Methodology and Evaluation

To forecast the cumulative COVID-19 cases, we use four different forecasting methods. Three of the forecasts are based on the autoregressive integrated moving average process which is usually denoted as *ARIMA* (*p,d,q*)where *p* is the order of the autoregressive model, *d* is the degree of differencing, and *q* is the order of the moving average model. The *ARIMA* model has been used for forecasting and assessing seasonality in infectious disease outbreaks.^12–15^

The *ARIMA* model is a generalization of the autoregressive moving average (*ARMA*) model with an ability to address the potential non-stationarity of the variable of interest. To test for stationarity of the cumulative COVID-19 cases, we used the Augmented Dickey-Fuller (*ADF*) and Phillips-Perron (*PP*) unit root tests. The null hypothesis of these tests is that the variable contains a unit root, hence non-stationary, whereas the alternative is that the time series variable was generated by a stationary process. Table 2 reports the p-values, for the unit root tests, for the 29 countries with the highest cumulative COVID-19 cases and at the aggregated level for the entire world. Table 2 in the Online Appendix provides the results of the unit root test for the 173 countries. The test results suggest non-stationarity which justifies the suitability of the *ARIMA* model.

**Table 2:** Results of the unit root tests for the 29 countries with the highest cumulative COVID-19 cases and at the aggregated-level of the entire world.

| Country | ADF | PP |
| --- | --- | --- |
| Bangladesh | 1.00 | 1.00 |
| Belarus | 0.98 | 0.98 |
| Belgium | 0.10 | 0.54 |
| Brazil | 1.00 | 1.00 |
| Canada | 0.23 | 0.79 |
| Chile | 1.00 | 1.00 |
| China | 1.00 | 0.99 |
| Ecuador | 0.57 | 0.56 |
| France | 0.26 | 0.63 |
| Germany | 0.43 | 0.72 |
| India | 1.00 | 1.00 |
| Iran | 0.01 | 0.17 |
| Italy | 0.52 | 0.75 |
| Mexico | 1.00 | 1.00 |
| Netherlands | 1.00 | 0.98 |
| Pakistan | 1.00 | 1.00 |
| Peru | 1.00 | 1.00 |
| Portugal | 0.97 | 0.93 |
| Qatar | 1.00 | 1.00 |
| Russia | 1.00 | 1.00 |
| Saudi Arabia | 1.00 | 1.00 |
| Singapore | 1.00 | 1.00 |
| South Africa | 1.00 | 1.00 |
| Spain | 0.82 | 0.84 |
| Sweden | 0.31 | 0.73 |
| Turkey | 0.97 | 0.92 |
| UAE | 1.00 | 1.00 |
| UK | 0.03 | 0.53 |
| USA | 0.42 | 0.86 |
| World | 1.00 | 1.00 |

Let *X*_*t*_ denote the cumulative COVID-19 cases on the *t*^th^ day for the country being analyzed. Then, *ARIMA* (*p,d,q*)process can be given as follows:

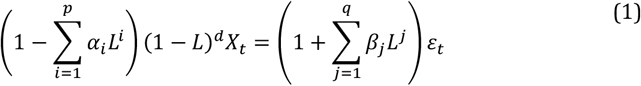

In equation (1), *L* is the lag operator, *α*’s and *β* ‘s are the coefficients of the autoregressive and the moving average component of the *ARIMA* model, respectively, and *ε* is the error term which is assumed to be independently and identically distributed from a normal distribution with zero mean. Equation (1) shows that the *AR* component allows the variable to be determined based on its prior values whereas the *MA* component shows that the error is a linear combination of the current and prior values of *ε*. The latter accounts for the autocorrelation in the variable of interest.

A particularly naïve attempt is to fit *ARIMA*(0,1,0)which is commonly referred to as random walk and its forecasts are termed as random walk forecasts (*RWF*). We generate the *RWF* with and without drift for the variable of interest.

A more systematic approach for fitting the *ARIMA* model follows these steps:^16,17^

1. To ensure stationarity, the differencing order (*d*) is selected by using the Kwiatkowski-Phillips-Schmidt-Shin (KPSS) test.
2. The lags, *p* and *q*, are determined by using the Akaike Information Criterion corrected for small sample sizes.

Aside from the *ARIMA* model, we also used the exponential smoothing method (*ETS*) for generating forecasts. *ETS* is a forecasting method for univariate data which deals with the systematic trend and seasonality, and can be used as an alternative to the *ARIMA* models.^18^

To evaluate the performance of forecasts, the data is divided into two mutually exclusive sets, the training and test sets. The training set is used to fit the model (without using any data from the test set) whereas the test set is kept for evaluating the forecast accuracy. We use a variant of the time series cross-validation which is a more sophisticated version of the usual training-test set methodology.^18^ In this method, there is a series of test sets, and each test set is accompanied by a corresponding training set consisting of observations before the test set. Therefore, a series of training-test sets are constructed, and for each training-test set forecast accuracy is determined. This method is more sophisticated than the usual training-test set methodology because it allows more comparisons of the forecasted and actual data values.

The time-series cross-validation method is also referred to as evaluation on a rolling forecasting origin because the origin of the test set is rolled forward in time. In simpler words:

1. An origin for the first test set is selected.
2. Forecasts are determined for the test set using the corresponding training set.
3. The origin is rolled forward by one period generating a new training-test set for which forecasts can be evaluated, and so on.

In this study, we take the 45^th^ day – since the first reported case in the country – as the origin which is then rolled forward one day at a time. The variation in our methodology is that, instead of taking each of the test set as a single observation, we take four different test sets for each training set: 1 week, 2 weeks, 3 weeks, and 4 weeks into the future. This allows us to ascertain the accuracy of the forecasting method up to 4 weeks ahead for each training set. Therefore, we include countries with at least 73 (45+28) observations to ensure that there is at least one available test set for the 4 weeks ahead forecasts for each country included for the forecast evaluation.

Suppose the country under consideration has data available for *t* ∈ {1,2 …, *T*}. The following steps explain the methodology:

1. Use the data available till *t* = 45, and forecast the values of *X*_*t*+*τ*_ for *τ* ∈ {1,2 …, 28} i.e. obtain the forecasts for the next 28 days or 4 weeks.
2. Construct 1 week, 2 weeks, 3 weeks, and 4 weeks ahead forecasts using the forecasted values till *t* + 7, *t* + 14, *t* + 21, and *t* + 28, respectively.
3. Increase the data sample by one day i.e. take the data till (*t* + 1)^th^ day and obtain 28-day ahead forecasts, and repeat this process until we reach the end of the data i.e., we reach the *T*^th^ day.

There are several methods to determine the accuracy of the forecasted values. We used the Mean Absolute Percentage Error (*MAPE*) for this purpose which is defined as follows:

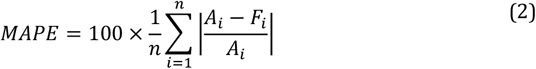

In equation (2), *A*_*i*_ and *F*_*i*_ denote the actual and forecasted values, respectively, and *n* is the number of forecasted values for which a corresponding actual data value exists. It should be clear that forecasting accuracy increases as *MAPE* becomes closer to zero. Since the forecasted variable of this study is the cumulative COVID-19 cases, *MAPE* represents the forecasting error as the percentage of cumulative COVID-19 cases. Based on our methodology, there is a series of training-test sets, and *MAPE* can be determined for each of the training-test set. Therefore, the forecasting accuracy is calculated by averaging *MAPE* over the series of the training-test sets.^18^

## Results

This section presents our results; Subsection 4.1 presents the forecasting accuracy of the four forecasting methodologies, and Subsection 4.2 presents the forecasted values and the heat maps.

### Forecasting Evaluation

Figure 1 shows the *MAPE* for the forecasted values for the 29 countries with the highest cumulative COVID-19 cases and at the aggregated level for the entire world. Figure 1 of the Online Appendix provides these results for the 173 countries. As expected, the *MAPE* increases as we increase the forecasting horizon from 1 week to 4 weeks ahead. This suggests that shorter-term forecasts are more accurate compared to longer-term forecasts. Overall, forecast evaluation shows that the *ARIMA* and *ETS* forecasts outperform *RWF* with and without drift.

**Figure 1:**
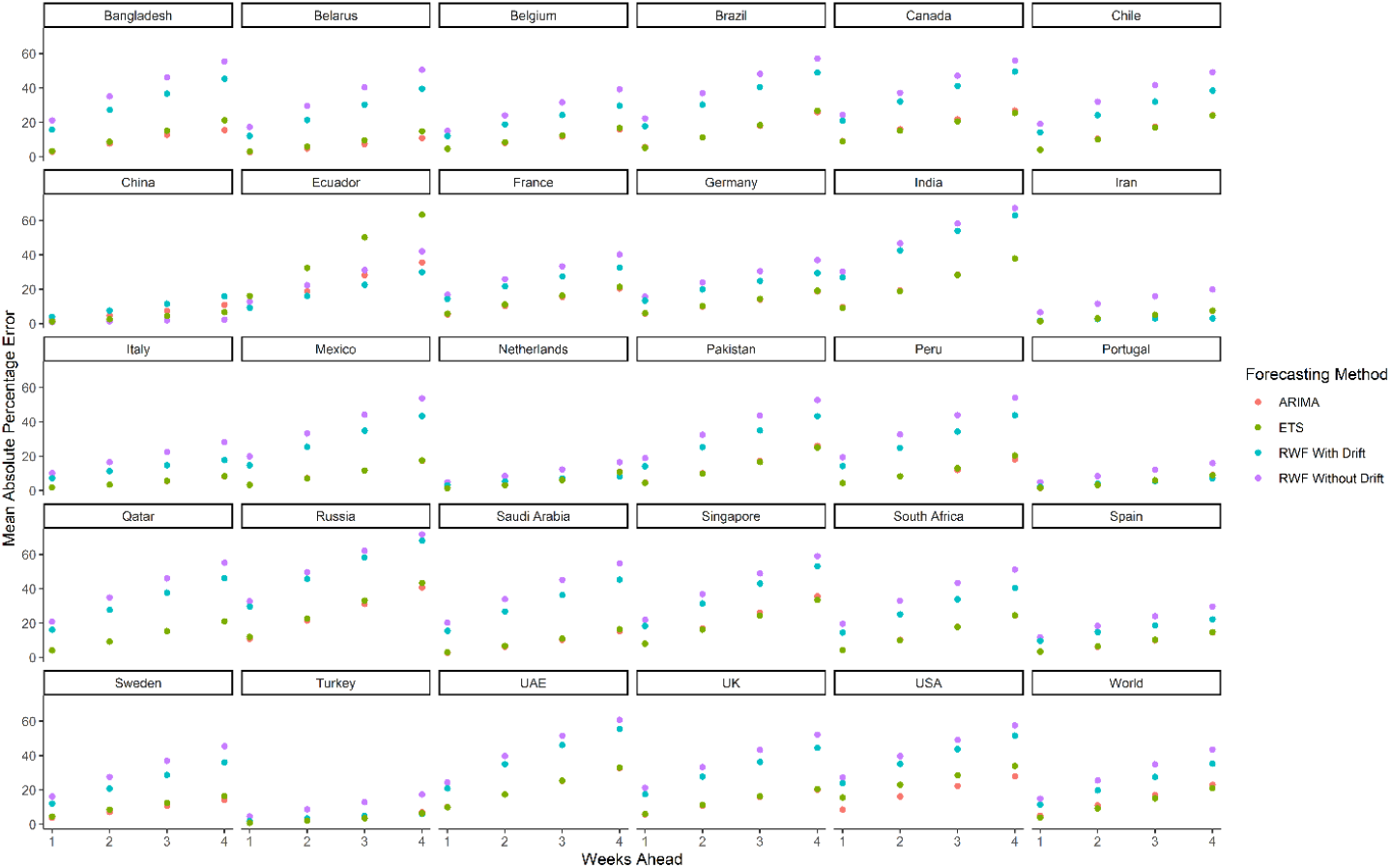
The *MAPE* for forecasted values of the 29 countries with the highest cumulative COVID-19 cases and at the aggregated level for the entire world.

Table 3 shows the average *MAPE* values for each forecasting horizon for the 173 countries and at the aggregated level for the entire world. In line with the observations of Figure 1, the *ARIMA* forecasts have low average *MAPE* values of 5.16%, 9.29%, 13.53%, and 18.20% for 1 week, 2 weeks, 3 weeks, and 4 weeks ahead, respectively. The *ETS* forecasts show similar average *MAPE* values as well. In comparison, *RWF* with drift has average *MAPE* values of 9.40% for 1 week, 15.92% for 2 weeks, 21.81% for 3 weeks, and 27.10% for 4 weeks ahead forecasts. *RWF* without drift exhibits even higher values than *RWF* with drift.

**Table 3:** Summary of the *MAPE* results for 173 countries and at the aggregated-level of the world.

| Weeks Ahead | Statistic | Forecasting Method |  |  |  |
| --- | --- | --- | --- | --- | --- |
|  |  | *ARIMA* | *ETS* | *RWF* With Drift | *RWF* Without Drift |
| 1 | Minimum | 0.09 | 0.00 | 1.54 | 0.04 |
|  | Maximum | 29.10 | 31.22 | 30.42 | 34.49 |
|  | Average | 5.16 | 5.28 | 9.40 | 11.23 |
|  | Std. Dev. | 4.85 | 5.39 | 6.63 | 8.64 |
| 2 | Minimum | 0.01 | 0.00 | 1.92 | 0.05 |
|  | Maximum | 42.85 | 42.17 | 48.81 | 53.84 |
|  | Average | 9.29 | 9.40 | 15.92 | 18.72 |
|  | Std. Dev. | 8.03 | 8.69 | 10.48 | 13.85 |
| 3 | Minimum | 0.02 | 0.00 | 2.25 | 0.06 |
|  | Maximum | 51.52 | 57.50 | 59.14 | 64.54 |
|  | Average | 13.53 | 13.67 | 21.81 | 25.00 |
|  | Std. Dev. | 10.55 | 11.67 | 13.39 | 17.76 |
| 4 | Minimum | 0.05 | 0.00 | 1.90 | 0.11 |
|  | Maximum | 64.33 | 67.39 | 68.00 | 71.52 |
|  | Average | 18.20 | 18.22 | 27.10 | 30.48 |
|  | Std. Dev. | 13.19 | 14.19 | 15.57 | 20.48 |

### Forecasted Scenario

Figure 2 shows the forecasted values generated using the *ARIMA* and *ETS* forecasting methodologies for the 29 countries with the highest cumulative COVID-19 cases and at the aggregated level for the entire world. Figure 2 of the Online Appendix provides these forecasts for the 173 countries.

**Figure 2:**
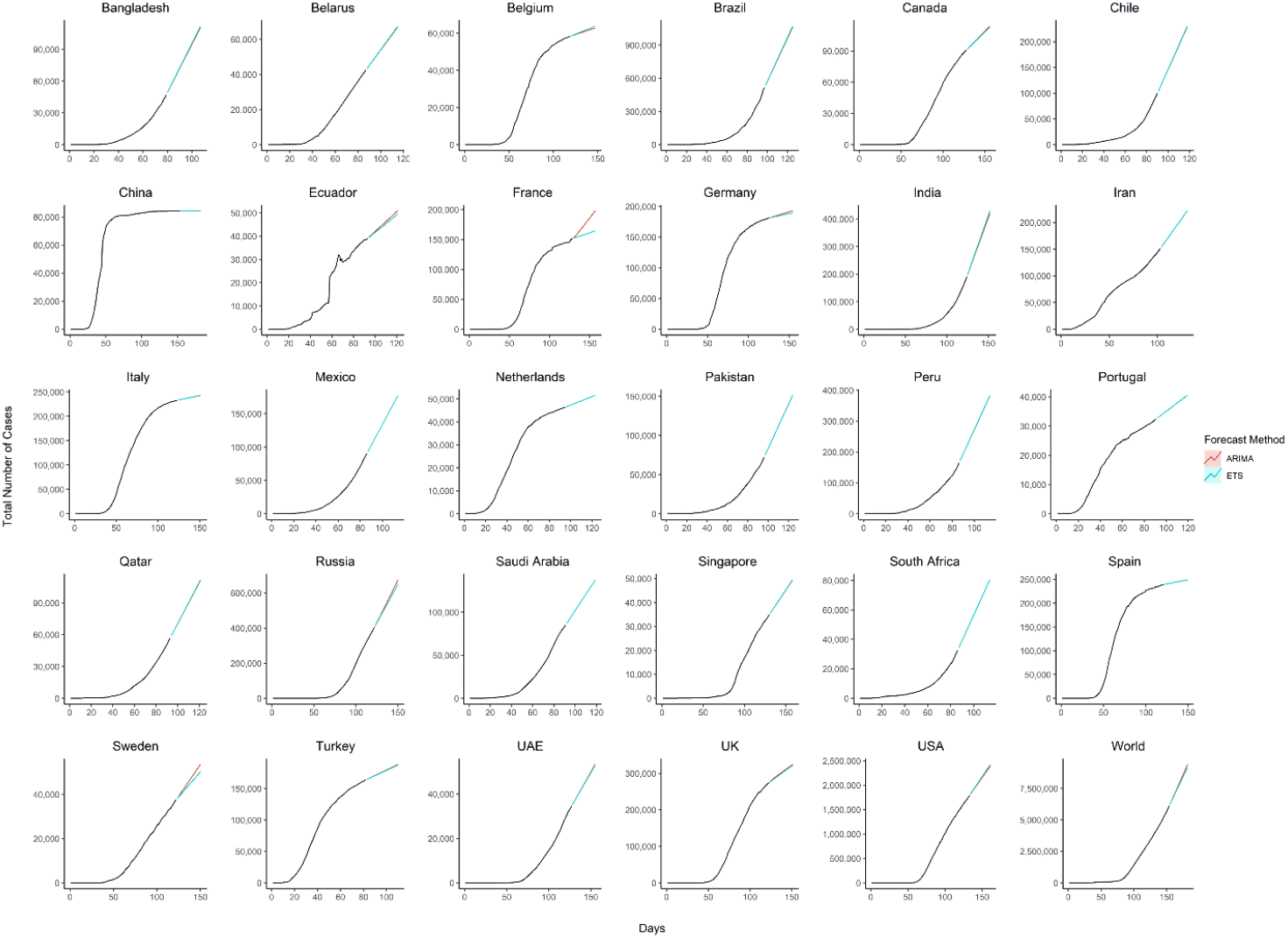
The forecasted values generated using the *ARIMA* and *ETS* forecasting methodologies for the 29 countries with the highest cumulative COVID-19 cases and at the aggregated level for the entire world.

We are only using the *ARIMA* and *ETS* forecasts because Subsection 4.1 showed that these forecasts outperform the *RWF* with and without drift. This figure uses the data from the entire sample period, and the values are forecasted for 4 weeks into the future. Figure 2 shows that the *ARIMA* and *ETS* forecasts perform similar except for a few cases. The figure shows the flattening of forecasts for Belgium, China, Germany, Italy, and Spain.

Figure 3 shows the heat maps based on the forecasted values of the cumulative cases of COVID-19. The graphs are generated using the forecasted values for 8^th^ June (Figure 3a), 15^th^ June (Figure 3b), 22^nd^ June (Figure 3c), and 29^th^June (Figure 3d).

**Figure 3:**
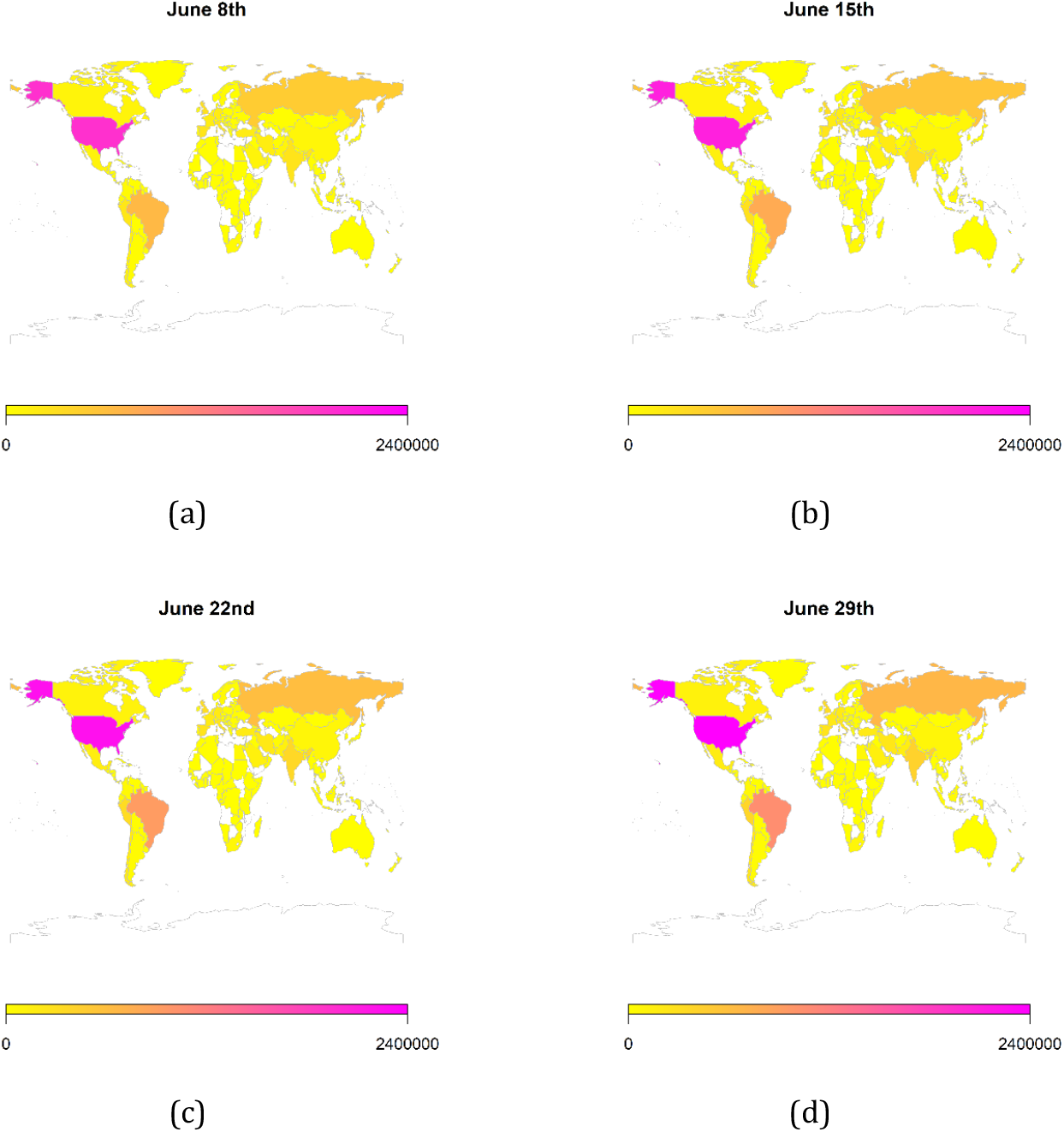
The heat maps generated using the forecasted values of the cumulative cases of COVID-19.

To generate the heat maps, we selected all those countries which had at least 73 (45+23) observations. Moreover, we only use the *ARIMA* forecasts here since its performance is comparable to the *ETS* forecasts. Overall, the heat maps depict information for 165 countries as some of the countries were not matched with the countries listed in the package used for generating these maps e.g. Palestine, Taiwan, etc. In Figure 3, during June the forecasted scenarios show an increase in the cumulative COVID-19 cases in South America, South Asia, and Russia.

## Discussion

Our results show that the *ARIMA* and *ETS* methods perform well in forecasting cumulative COVID-19 cases. Additionally, using these forecasts, we generated heat maps to provide a pictorial representation of the countries at risk of having an increase in cases in the coming 4 weeks in June.

Globally, uncertainty exists around the spread and transmissions of SARS-CoV2. For this purpose, many mathematical modeling and simulation-based techniques, especially compartmental model techniques, to better understand the transmissions of COVID-19 cases. Among these, the most used is the Susceptible-Exposed-Infectious-Recovered (SEIR) model.^19–22^ The SEIR model makes assumptions on the population belonging to the different compartments. However, for these assumptions to be reliable, large datasets are required.^19–22^ During a pandemic, not a lot of data is available to reliably run the aforementioned models. In comparison, the data-driven methods considered in this paper are less data-hungry and do not require as much level of detail in the datasets for forecasting purposes. Other advantages of these data-driven techniques include simplicity of estimation which can be performed using open-source statistical software (RStudio).

Different countries and regions have different health systems and capacities in place which determine their testing capabilities. The SEIR model can capture the propagation of the disease which means that it would be able to predict the true number of cases considering the susceptible and asymptomatic individuals. However, data for asymptomatic cases is largely unavailable for SARS-CoV2 due to limited testing capabilities and a large proportion of asymptomatic cases not being detected; making it challenging to verify the predictions from the compartmental models. On the other hand, data-driven techniques can provide information on the confirmed number of cases with high accuracy. For this study we focused on the cumulative COVID-19 cases, however, the forecasting methods can be used for other indicators such as cumulative deaths, cumulative recovery, etc. The forecasts of the confirmed number of cases are sensitive to the number of tests performed, however, since the confirmed number of cases is an indicator of the anticipated burden on the healthcare system and professionals, the projections by the data-driven techniques might be insightful for the policymakers. This is important because the availability of health service resources during COVID-19 is an issue faced by many countries. One study carried out in different states of the United States predicted an excess demand of 64,175 total beds and 17,380 ICU beds at the peak of COVID-19.^23^ Even with lockdown measures enacted, the peak demand for healthcare services, during the COVID-19 pandemic, is likely to exceed capacity irrespective to the current capability of the healthcare infrastructure and resources.

Globally, our forecasting results reveal that the number of cases will increase in most of the countries. Forecasted scenarios for June depict that some of the hardest-hit regions include the United States, the United Kingdom, Canada, some parts of Europe, South America, and South Asia as shown in Figure 2 of the Online Appendix. Middle eastern countries including Iran, the United Arab Emirates, and Saudi Arabia also show an increase in the cumulative COVID-19 cases.

The future of the global pandemic greatly depends on the implementation of mitigation measures. Strict measures such as worldwide lockdowns, travel restrictions, school closures, non-essential business closures, social distancing, isolation of infected populations as well as heightened hygiene measures can potentially reduce the risk of spread.^23^ The forecasted results, Figure 2 of the Online Appendix, of the following countries show a flattening of the curve possibly due to timely containment and mitigation measures: Andorra, Antigua and Barbuda, Aruba, Australia, Austria, Barbados, Belgium, Brunei, China, Croatia, Cyprus, Faeroe Islands, French Polynesia, Germany, Greece, Guernsey, Iceland, Isle of Man, Italy, Latvia, Liechtenstein, Luxembourg, Mauritius, Montenegro, New Zealand, Norway, San Marino, Seychelles, Slovakia, Slovenia, South Korea, Spain, Switzerland, Taiwan, Thailand, Trinidad and Tobago, and Vietnam. However, the effectiveness of interventions is far from homogenous and depends on how well people comply, the presence of enforcement, how well testing/contact tracing/quarantine efforts that are run alongside the lockdown are performed, etc. Yet, hopes of curtailing the pandemic have proven elusive, with many countries forced by their economies to relax the quarantine measures which can potentially lead to an exponential increase in the number of cases. A warning has also been issued by the WHO that a spike in the number of cases might be observed as countries are relaxing the lockdown measures which may lead to a potential second wave of the COVID-19.

Although, the novel coronavirus pandemic is associated with many uncertainties, we believe that forecasting and predictive modeling can be an effective tool in targeted intervention strategies. Model-based predictions can help policymakers to make the right decisions in a timely way.^24^

## Conclusion

Our results indicate that the *ARIMA* and *ETS* model perform well in forecasting the cumulative COVID-19 cases. We ran the model for 173 countries with varying health system resources and infrastructure, and at the aggregated level for the entire world. The results suggest that the *ARIMA* and *ETS* model can be used for SARS-CoV2 forecasting in different countries and regions with a high level of accuracy. Since these models only rely on past observations of the cumulative 0COVID-19 cases, it can also be used for forecasting provincial, district, or state level SARS-CoV2 cases and other COVID-19 indicators.

## Data Availability

The data was acquired from ourworldindata.org which is publicly accessible online.

---

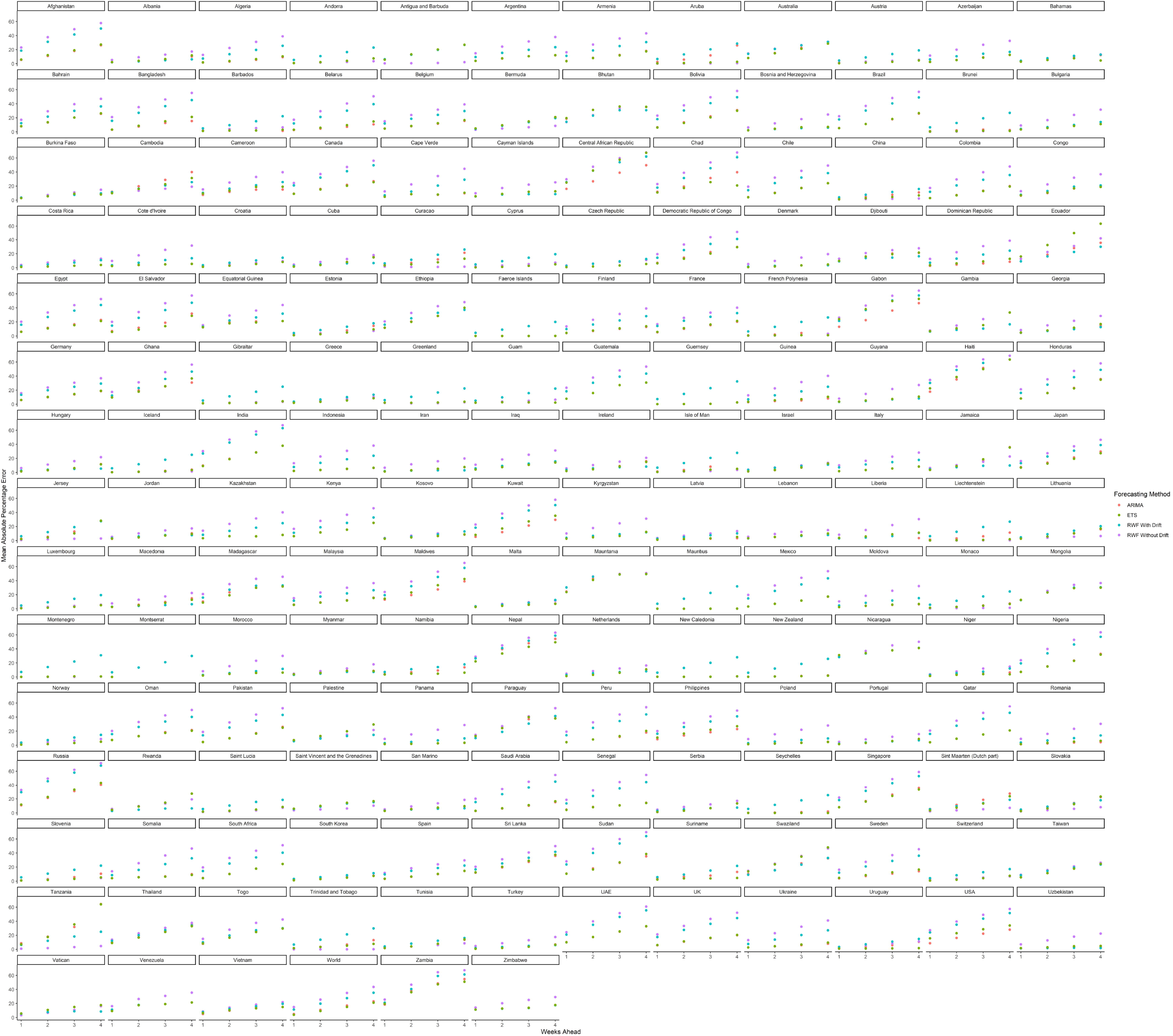

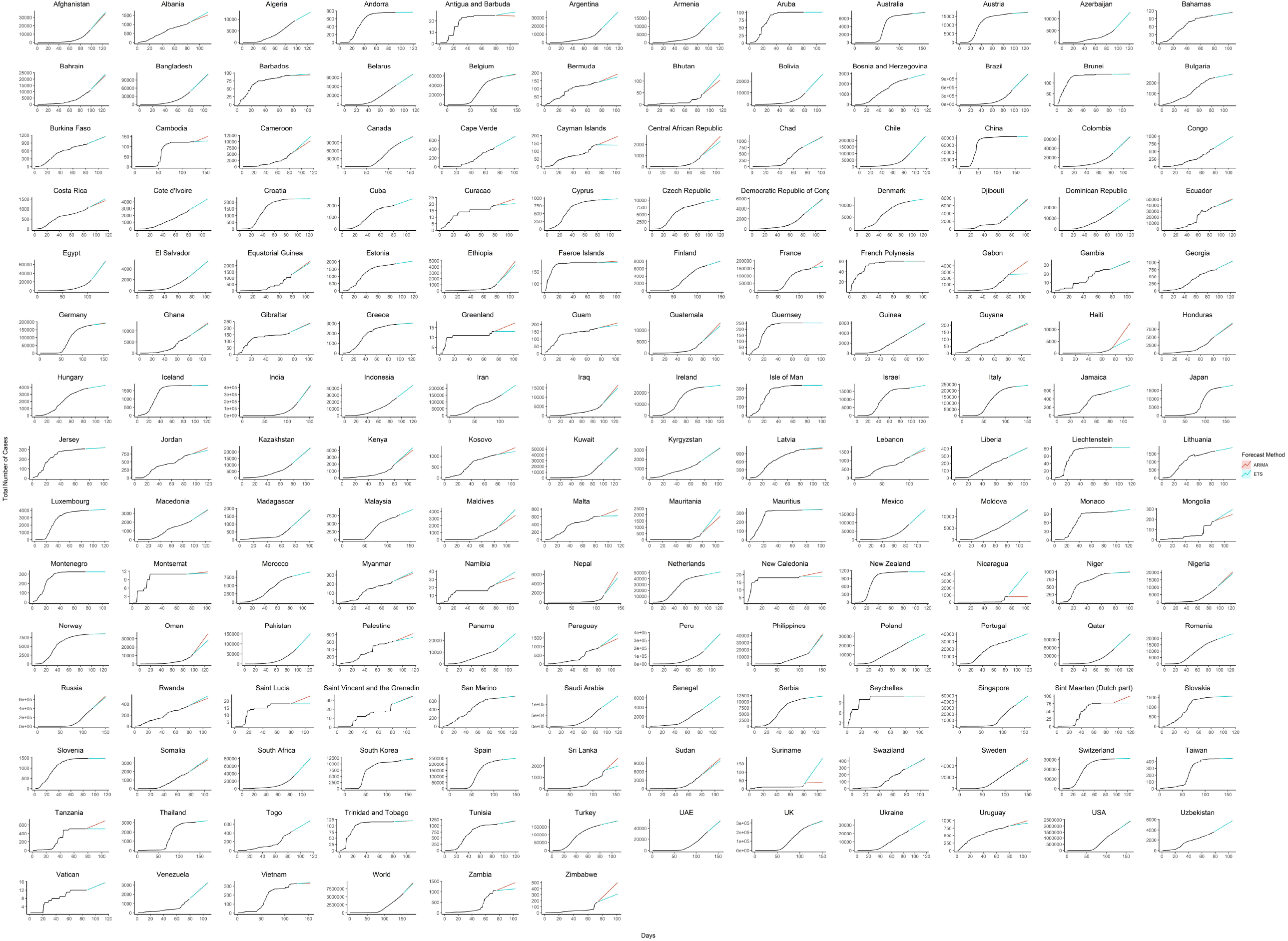

